# Causal Effects of Physical Activity and Sedentary Behavior on Healthcare Costs

**DOI:** 10.64898/2026.08.18.26360577

**Authors:** Eliisa Mäkelä, Jaana T. Kari, Sander Van Genechten, Reijo Bottas, Elina Sillanpää, Laura Joensuu

**Author notes:** Corresponding author: Eliisa Mäkelä, M.Sc.

## Abstract

**Importance:** While increased physical activity (PA) and decreased sedentary behavior (SB) are associated with favorable health outcomes, evidence regarding their causal effects on healthcare costs remains limited.

**Objective:** To assess the causal effects of PA and SB on healthcare costs.

**Design:** A two-sample Mendelian randomization (MR) study.

**Setting:** Separate, non-overlapping cohorts with genetic instruments for self-reported and device-based PA and SB, and healthcare costs.

**Participants:** The instruments used to assess self-reported PA were derived from a genome-wide meta-analysis of 606,820 individuals across 51 cohorts. Two large genome-wide association studies (GWASs) were used for self-reported SB (leisure screen time N=526,725; television watching N=408,815), while accelerometer-based GWASs (N=89,683–91,105) were used for device-based PA and SB. The instruments used to assess the outcome data were obtained from the FinnGen cohort (N=373,160).

**Exposures:** Genetically predicted PA and SB.

**Main Outcomes and Measures:** Validated genetic instruments for log-transformed annual healthcare costs derived from registers, including primary care, secondary care, and medication costs. Inverse variance weighting was used as the primary MR measure, while the sensitivity analyses included MR-Egger, weighted median, simple mode, weighted mode, F-score, Cochran’s Q, and leave-one-out analysis.

**Results:** Higher genetically predicted self-reported PA was associated with lower healthcare costs (causal estimate, β=-0.166; 95% CI, -0.270 to -0.062). In contrast, higher genetically predicted SB (leisure screen time or television watching) was associated with higher healthcare costs across self-reported datasets (β=0.097; 95% CI, 0.064 to 0.130; β=0.114; 95% CI, 0.063 to 0.165, respectively). No associations were observed for device-based PA (β=-0.014; 95% CI, -0.040 to 0.014) or SB (β=-0.009; 95% CI, -0.197 to 0.179).

**Conclusions and Relevance:** Findings based on genetically predicted PA and SB support a causal association between these behaviors and healthcare costs, suggesting that increasing population’s leisure-time PA and reducing SB may decrease healthcare expenditure. This highlights the importance of promoting PA for both population health and long-term sustainability of healthcare systems. However, causal evidence remains partly limited, particularly for device-based measures of these behaviors.

**Key Points:** *Question:* Are physical activity and sedentary behavior causally associated with healthcare costs?

*Findings:* In this two-sample Mendelian randomization study, higher genetically predicted self-reported physical activity was associated with lower annual healthcare costs, whereas higher genetically predicted self-reported sedentary behavior was associated with higher annual healthcare costs. No associations were observed for device-based measures of either physical activity or sedentary behavior.

*Meaning:* These findings support potential causal effects of physical activity and sedentary behavior on healthcare expenditure, suggesting that policies and interventions aimed at increasing leisure-time physical activity and reducing sedentary time may reduce healthcare costs while improving population health.

## Introduction

Physical inactivity, which is defined as not meeting current physical activity guidelines,^1^ and sedentary behavior, as characterized by waking behavior with an energy expenditure of ≤1.5 metabolic equivalents of task (METs) while sitting, reclining, or lying down,^2^ are associated with increased risks of morbidity and mortality^3–5^ and with a substantial economic burden.^6,7^ This encompasses direct healthcare expenditures and indirect costs related to productivity and labor market outcomes.^6,8–10^ In terms of direct costs, research suggests that physical inactivity is associated with approximately 0.3–5% higher national healthcare expenditure,^6^ while individual-level healthcare costs are 9–27% lower for physically active individuals than for those with low levels of physical activity.^10^ While relatively few studies have examined the economic burden of sedentary behavior, prolonged sedentary time is associated with higher healthcare costs.^7,8,11,12^ However, estimates concerning the general population have largely been derived from observational data and using methods such as population-attributable fractions,^6^ meaning that they provide limited support for causal inference.

Establishing the causal effects of physical activity and sedentary behavior on healthcare costs is challenging, largely due to the limited feasibility of conducting randomized controlled trials with sufficiently long intervention periods.^13^ While recent Mendelian randomization studies have strengthened the evidence of the causal links between physical activity, sedentary behavior and diverse health outcomes,^14–16^ evidence of their causal effects on healthcare costs remains limited. Yet robust and policy-relevant cost estimates are essential for assessing economic impacts and informing decision-making.

The present study addresses this gap in the literature by applying a genetically informed two-sample Mendelian randomization (2SMR) approach to estimate the causal effects of physical activity and sedentary behavior on healthcare costs. We hypothesized that higher physical activity would be associated with lower healthcare costs, whereas higher sedentary behavior would be associated with higher healthcare costs.

## Methods

We applied a 2SMR approach using genetic variants as instrumental variables (IVs). More specifically, 2SMR identifies genetic variants that are robustly associated with the exposure (i.e., relevance assumption) and then estimates the association between the IVs and the outcome. This approach relies on the core assumptions that these genetic variants are independent of confounders and only affect the outcome through the exposure (i.e., independence and exclusion restriction assumptions, respectively).^17^ In addition, the genetic variants are assumed to be randomly allocated at the point of conception, approximating a randomized controlled trial (RCT) and supporting causal inference in cases where a RCT is neither ethical nor feasible. Here, the exposure is either physical activity or sedentary behavior, as measured using both self-reported and device-based metrics, while the outcome is register-based annual healthcare costs. We utilized IVs from publicly available genome-wide association studies (GWASs) conducted in cohorts approved by the respective ethical boards. Therefore, no additional ethical approval was required. The reporting of this study follows the Strengthening the Reporting of Observational Studies in Epidemiology Using Mendelian Randomization (STROBE-MR) guidelines.^18^

### Data Sources and Instruments

We reviewed published GWASs concerning physical activity and sedentary behavior and selected those with the largest number of participants at the time of analysis and/or with sufficient statistically significant single-nucleotide polymorphisms (SNPs) at the genome-wide level (typically p < 5 × 10^-8^) to ensure adequate statistical power (Table 1). Moreover, we used non-overlapping GWASs for the exposures and outcomes, all of which were based on participants with European ancestry.

**Table 1.**
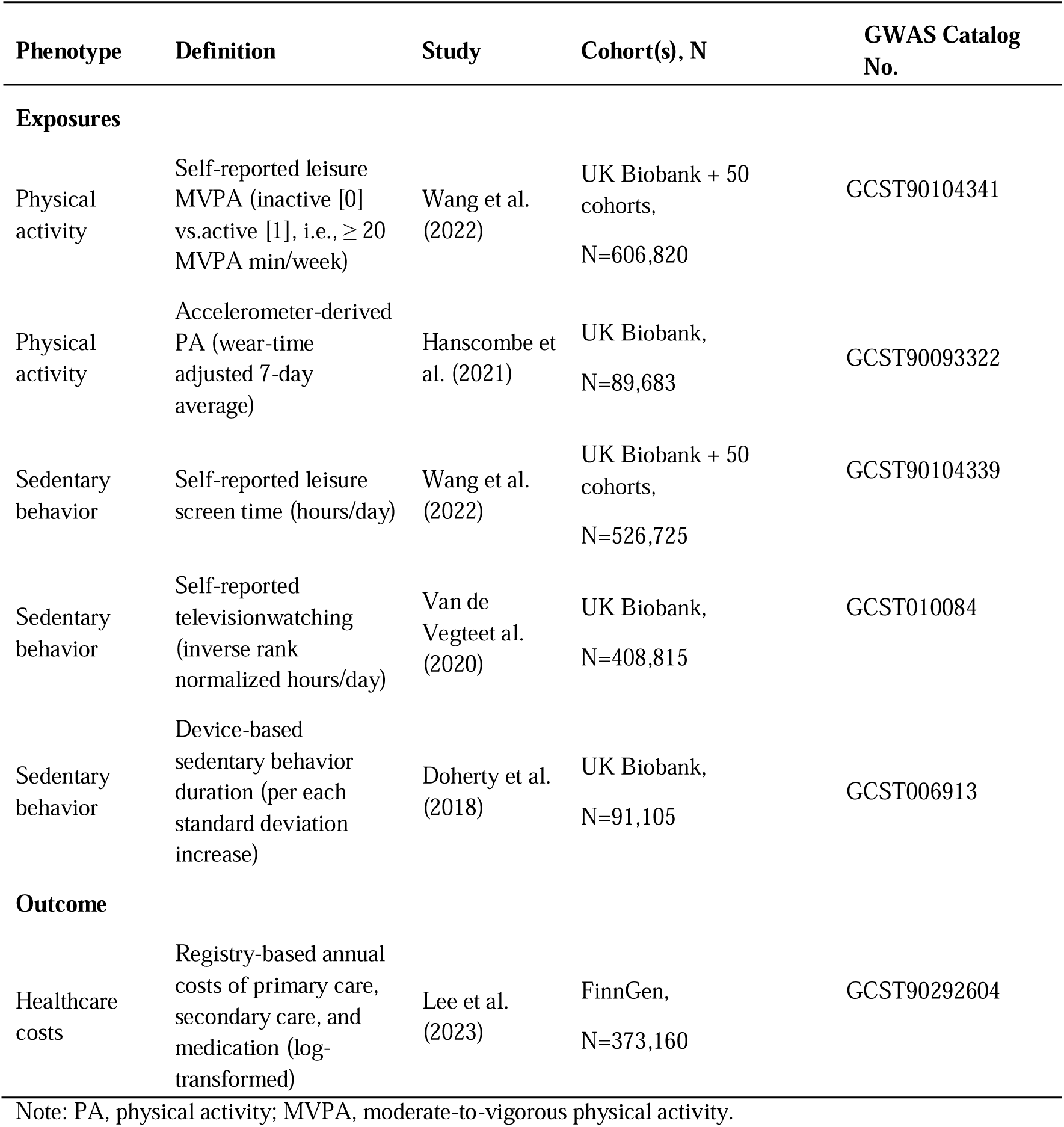
Characteristics of the Data Sources.

| Phenotype | Definition | Study | Cohort(s), N | GWAS Catalog No. |
| --- | --- | --- | --- | --- |
| **Exposures** |  |  |  |  |
| Physical activity | Self-reported leisure MVPA (inactive [0] vs. active [1], i.e., $\geq 20$ MVPA min/week) | Wang et al. (2022) | UK Biobank + 50 cohorts, N=606,820 | GCST90104341 |
| Physical activity | Accelerometer-derived PA (wear-time adjusted 7-day average) | Hanscombe et al. (2021) | UK Biobank, N=89,683 | GCST90093322 |
| Sedentary behavior | Self-reported leisure screen time (hours/day) | Wang et al. (2022) | UK Biobank + 50 cohorts, N=526,725 | GCST90104339 |
| Sedentary behavior | Self-reported television watching (inverse rank normalized hours/day) | Van de Vegteet al. (2020) | UK Biobank, N=408,815 | GCST010084 |
| Sedentary behavior | Device-based sedentary behavior duration (per each standard deviation increase) | Doherty et al. (2018) | UK Biobank, N=91,105 | GCST006913 |
| **Outcome** |  |  |  |  |
| Healthcare costs | Registry-based annual costs of primary care, secondary care, and medication (log-transformed) | Lee et al. (2023) | FinnGen, N=373,160 | GCST90292604 |

Note: PA, physical activity; MVPA, moderate-to-vigorous physical activity.

### Exposures

#### Physical Activity

The IVs for self-reported physical activity were derived from a dataset comprising 51 cohorts, which included UK Biobank and external cohorts consisting of a total of 606,820 individuals (Table 1).^19^ The variable of interest here describes individuals being active or inactive based on the reported volumes of leisure-time moderate-to-vigorous physical activity (MVPA) harmonized across cohorts in a recent meta-analysis.^19^ Owing to the zero-inflated distribution, MVPA was dichotomized as inactive (0) versus active (1) using a threshold of ≥20 minutes per week (i.e., approximately the cohort median). We used UK Biobank participants with available accelerometer data (N=89,683) for the device-based physical activity IVs.^20^ This phenotype describes the wear-time adjusted seven-day average physical activity. Device-based physical activity GWASs are generally limited when it comes to statistically significant SNPs,^21–23^ thus the selected GWAS with nine reported SNPs provided sufficient utility to conduct MR.

#### Sedentary Behavior

We used three GWAS datasets for sedentary behavior—namely, two self-reported measures and one device-based measure (Table 1). The first dataset included 526,725 individuals and used leisure screen time as a measure of sedentary behavior.^19^ To address concerns regarding potential pleiotropy observed in the first analysis, we used a second dataset involving self-reported measures,^24^ which included 408,815 individuals of European ancestry and genetic variants associated with television watching. The third dataset included 91,105 individuals with accelerometer-derived data concerning sedentary behavior (seven-day period).^23^ Here, the researchers trained a machine learning model to predict sedentary behaviors (generally ≤1.5 METs) from wrist-worn accelerometry data. The variable indicates the probability to engaging in a sedentary behavior across the assessed time window and is expressed as the standard deviation in the duration of the behavior.^23^

### Outcome

Healthcare costs were obtained from the FinnGen study, which involved 373,160 individuals.^25^ More specifically, FinnGen is an ongoing Finnish public–private partnership research project that links genotype and health registry data using personal identification numbers and currently includes data from nearly 10% of the Finnish population.^26^ The healthcare costs phenotype is a log-transformed measure of the total annual costs, including primary care, secondary care, and prescription medication costs, adjusted for each individual’s follow-up time in the respective registry. The follow-up period extended from 1998 for the secondary care and prescription medication registries and from 2011 for the primary care registry through to the end of follow-up or 2021 at the latest.^25^ Costs were calculated based on the average costs of Finland’s healthcare system, standardized to the 2017 euro values, according to the service type (e.g., outpatient care, home care, mental healthcare, etc.), specialty (e.g., internal medicine, emergency medicine, surgery, etc.), and visit type (e.g., inpatient ward, emergency care, other). In the FinnGen study, the researchers assessed the robustness of the genetic instruments and observed the genetic correlations between Finland, United Kingdom, and the Netherlands (all r_g_>0.077, p<.001), indicating the cross-country comparability.^25^ The genotyping procedures and covariates used for all the GWASs are detailed in the Supplement.

### Statistical Analysis

We used the TwoSampleMR R package^27^ to estimate the causal effects of each exposure on the log-transformed annual healthcare costs, as expressed per one-unit increase in the exposure. Following standard MR procedures, SNPs from each exposure GWAS were clumped within a 10,000-kilobase window using a linkage disequilibrium threshold of r2=0.001. The SNP with the lowest P-value in each clump was retained as the representative SNP. The inverse variance weighted (IVW) method was prespecified as the primary causal estimate. We assessed the compliance with the main MR assumptions (i.e., relevance, independence, and exclusion restriction) and the robustness of the findings using additional methods, including F-statistics, weighted median, MR-Egger, simple mode, weighted mode, MR-Egger intercept, Cochran’s Q statistics, and leave-one-out analyses.^17^ Statistical significance was defined as p<0.05. The relevance of our findings was also considered in relation to approximated population-level cost estimates based on European Union (EU) health expenditure per capita obtained from the World Bank, the prevalence of physical inactivity obtained from the Organisation for Economic Co-operation and Development (OECD), and European population statistics obtained from Eurostat.^28–30^

## Results

### Physical Activity

The IVW analysis indicated that higher genetically predicted physical activity was associated with lower healthcare costs (IVW β=-0.166; 95% confidence interval [CI] -0.270, -0.062; Table 2). This estimate corresponds to approximately 15.3% (95% CI, 6.0% to 23.7%) lower annual healthcare for a theoretical 100 percentage-point increase in the prevalence of being physically active, equivalent to an approximately 1.5% lower expenditure for a 10 percentage-point increase in prevalence.^31^ The weighted median (β=-0.084; 95% CI, -0.170 to 0.002), simple mode (β=-0.086; 95% CI, -0.225 to 0.053), and weighted mode (β=-0.072; 95% CI, -0.180 to 0.036) estimates showed effects in the same direction as the IVW estimate, whereas the MR-Egger estimate (β=0.049; 95% CI, -0.552 to 0.650) was imprecise and close to null. Overall, the sensitivity analyses were directionally consistent with the IVW estimate, albeit only the IVW estimate reached statistical significance. The analysis was based on 14 SNPs with F-scores > 30. A scatter plot of the relevant data is available in eFigure 1. The MR-Egger intercept test (0.049, p=0.552) showed no evidence of directional pleiotropy, while the leave-one-out analysis did not identify influential outlier SNPs (eFigure 2).

**Table 2.** Associations of Genetically Predicted Physical Activity and Sedentary Behavior with Healthcare Costs.

| Causal estimates | Method | nSNP | $\beta$ (SE) | 95% CI |
| --- | --- | --- | --- | --- |
| **Physical activity** |  |  |  |  |
| Self-reported leisure MVPA, active vs. inactive (Wang et al. 2022) | IVW | 14 | **-0.166 (0.053)** | **-0.270, -0.062** |
|  | MR-Egger | 14 | 0.049 (0.307) | -0.552, 0.650 |
|  | Weighted median | 14 | -0.084 (0.044) | -0.170, 0.002 |
|  | Simple mode | 14 | -0.086 (0.071) | -0.225, 0.053 |
|  | Weighted mode | 14 | -0.072 (0.055) | -0.180, 0.036 |
| Accelerometer-derived 7-day average (Hanscombe et al. 2021) | IVW | 3 | -0.014 (0.014) | -0.041, 0.014 |
|  | MR-Egger | 3 | -0.019 (0.097) | -0.209, 0.017 |
|  | Weighted median | 3 | -0.013 (0.010) | -0.033, 0.007 |
|  | Simple mode | 3 | -0.013 (0.016) | -0.044, 0.017 |
|  | Weighted mode | 3 | -0.015 (0.013) | -0.040, 0.010 |
| **Sedentary behavior** |  |  |  |  |
| Self-reported leisure screen time, h/day (Wang et al. 2022) | IVW | 104 | **0.097 (0.017)** | **0.064, 0.130** |
|  | MR-Egger | 104 | -0.100 (0.076) | -0.249, 0.049 |
|  | Weighted median | 104 | **0.085 (0.018)** | **0.050, 0.120** |
|  | Simple mode | 104 | 0.090 (0.060) | -0.028, 0.208 |
|  | Weighted mode | 104 | 0.077 (0.063) | -0.046, 0.200 |
| Self-reported television watching, h/day (van de Vegte et al. 2020) | IVW | 92 | **0.114 (0.026)** | **0.063, 0.165** |
|  | MR-Egger | 92 | 0.144 (0.130) | -0.111, 0.399 |
|  | Weighted median | 92 | **0.120 (0.028)** | **0.065, 0.175** |
|  | Simple mode | 92 | 0.151 (0.098) | -0.041, 0.343 |
|  | Weighted mode | 92 | 0.137 (0.111) | -0.081, 0.355 |
| Accelerometer-derived sedentary<br>behavior duration, per 1 SD (Doherty<br>et al. 2018) | IVW | 3 | -0.009 (0.096) | -0.197, 0.179 |
|  | MR-Egger | 3 | -0.268 (0.155) | -0.572, 0.036 |
|  | Weighted median | 3 | -0.042 (0.085) | -0.209, 0.125 |
|  | Simple mode | 3 | -0.031 (0.132) | -0.290, 0.228 |
|  | Weighted mode | 3 | -0.072 (0.175) | -0.415, 0.271 |

Values are causal estimates ( $\beta$ ), standard errors (SE), and 95% confidence intervals (95% CI); IVW, inverse variance weighted; MR, Mendelian randomization; nSNP, number of single-nucleotide polymorphisms used for modeling; 95% CI, 95% confidence interval; SD, standard deviation; statistically significant associations are highlighted in bold.

No statistically significant associations were observed for device-based physical activity (three SNPs with F-scores > 27) (Table 2, eFigure 3 and 4). A more relaxed instrument selection threshold (P < 5 × 10^-7^) increased the number of SNPs to 13 without changing the overall interpretation.

### Sedentary Behavior

The IVW analysis based on 104 SNPs (F-scores > 30) revealed a significant positive association between genetically predicted leisure screen time and healthcare costs (β=0.097; 95% CI, 0.064 to 0.130), corresponding to an estimated 10.2% (95% CI, 6.6% to 13.9%) increase in annual healthcare costs per one hour change in daily leisure screen time (Table 2). Similar estimates were observed when using the weighted median method (β=0.085; 95% CI, 0.050 to 0.120), whereas the MR-Egger and mode-based estimates were not statistically significant (see eFigure 5 for the scatter plot). The leave-one-out analysis showed stable estimates after sequential exclusion of individual SNPs (eFigure 6). Yet significant heterogeneity was observed in the IVW model (Q=275.4, p<0.001), and the MR-egger intercept (0.005, p=0.009) showed evidence of directional horizontal pleiotropy.

To assess the robustness of these findings, we conducted an additional analysis using the GWAS concerning television watching.^24^ The IVW analysis based on 92 SNPs (F-scores > 17) indicated a significant positive association between higher genetically predicted sedentary behavior and annual healthcare costs (β=0.114; 95% CI, 0.063 to 0.165), corresponding to a 12.1% (95% CI, 6.5% to 17.9%) increase in annual healthcare costs. The causal estimates were directionally consistent between the two self-reported sedentary behavior datasets (see the scatter plot in eFigure 7 and the leave-one-out analysis eFigure 8). However, no evidence of directional horizontal pleiotropy was observed for television watching (MR-Egger intercept -0.001, p=0.81).

For device-based sedentary behavior, relaxing the genome-wide significance threshold to p < 5 × 10^-4^ yielded three SNPs after harmonization. The IVW analysis showed no association with annual healthcare costs (β=-0.009; 95% CI, -0.197 to 0.179; see eFigure 9 for the scatter plot and eFigure 10 for the leave-one-out analysis). Interpretation of the device-based findings is limited by the low number of genetic instruments.

## Discussion

In this 2SMR study, we estimated the causal effects of physical activity and sedentary behavior on healthcare costs. We found higher genetically predicted physical activity to be associated with lower annual healthcare costs (-0.166; 95% CI, -0.270 to -0.062), whereas higher genetically predicted sedentary behaviors were associated with higher annual healthcare costs (0.097; 95% CI, 0.064 to 0.130 for leisure screen time and 0.114; 95% CI, 0.063 to 0.165 for television watching). These findings corresponded to approximately 15% lower annual health care costs among individuals reporting at least 20 minutes of leisure physical activity per week, whereas a 1-hour increase in daily leisure screen time or television watching was associated with approximately 10% and 12% higher costs, respectively. Causal associations were observed for the self-reported measures but not for the device-based measures of physical activity and sedentary behavior.

Healthcare spending is among the fastest-growing categories of expenditure worldwide and represents a major policy concern for governments and healthcare systems. Indeed, in 2022, global healthcare expenditure reached US$10 trillion, accounting for 9.9% of the global gross domestic product (GDP).^32^ Given the well-established adverse health outcomes associated with physical inactivity and sedentary behavior,^3^ as well as the persistently low levels of physical activity worldwide,^34^ research interest in the economic consequences of physical activity has grown in recent years. Our findings extend earlier research by providing evidence that leisure-time physical activity and sedentary behavior may causally influence healthcare costs, and suggests that even modest increases in physical activity and reductions in daily leisure screen time in the population may have meaningful implications for annual healthcare expenditure.

Based on our estimates, if the one-third of EU adults who reported engaging in no leisure-time physical activity^29^ increased their activity levels, healthcare costs could potentially be reduced by approximately 5.3%. This corresponds to savings of approximately €350 per previously inactive individual, or roughly €43 billion overall,^30^ based on the average annual healthcare expenditure in the EU in 2024.^28^ By leveraging the MR approach, this study provides evidence that is less susceptible to the confounding and reverse causation that may have affected prior observational studies, thereby strengthening the evidence of a causal relation between physical activity and sedentary behavior and healthcare costs. However, these estimates are illustrative and assume that the MR effect scales linearly at the population level.

The present findings are supported by biological evidence. Previous studies have found that regular physical activity preserves mitochondrial function, reduces chronic low-grade inflammation, and improves metabolic homeostasis, whereas prolonged physical inactivity impairs these health-protective processes.^36–38^ Together with the epidemiological evidence of adverse health outcomes being associated with sedentary behavior and physical inactivity,^3,39,40^ these biological effects provide a plausible mechanism linking physical activity and sedentary behavior to chronic diseases and, ultimately, to healthcare utilization and costs. Although physical activity may also be associated with adverse health outcomes, including injuries requiring medical attention and increased healthcare utilization,^33,41^ our findings suggest that the overall health and economic benefits of physical activity outweigh these potential harms.

As physical activity is often viewed primarily as an individual health responsibility, its broader societal and economic benefits may be under-represented in policy discussions.^35^ Our findings reinforce the importance of promoting physical activity and reducing sedentary behavior as public health priorities with the potential to generate economic benefits. These findings further suggest that even modest increases in physical activity can translate into substantial reductions in healthcare costs, which is consistent with recent evidence indicating that an additional five minutes per day of MVPA may yield meaningful population-level health benefits.^42^ Additionally, the observed economic estimates for sedentary behavior (10–12%) are comparable in magnitude to those reported for well-established clinical risk factors, including body mass index and systolic blood pressure, for which a one standard deviation increase has been associated with 13–14% higher annual healthcare costs.^25^

Yet data on public investments aimed at reducing sedentary behavior are limited. Estimates on government expenditures on recreational and sporting services, as well as on primary prevention, indicate that public investments in these areas are of a similar magnitude, although each accounted for less than 1% of the total general government expenditure or <0.5% of GDP in the EU in 2023.^43,44^ The findings of this study suggest that greater priority should be given to preventive measures in the future, including promoting physical activity and reducing sedentary behavior as key strategies for improving population health and reducing healthcare costs.

In contrast to the findings concerning self-reported measures, no clear associations were observed for the device-based measures of either physical activity or sedentary behavior. Several factors may account for this discrepancy. While self-reported measures are susceptible to recall bias and measurement error,^45^ and may be influenced by subjective perceptions of health, they are generally derived from larger cohorts with more participants, greater genetic variability, and consequently, higher statistical power. Conversely, device-based measures provide a more objective assessment of movement and sedentary time,^46^ but generally do not distinguish between behavioral domains such as leisure-time and occupational physical activity, which may have different associations with health.^47,48^

### Limitations

This study has several limitations. First, the device-based GWASs included fewer genetic instruments, limiting statistical power and precision. Second, heterogeneity and pleiotropy were observed in the sedentary behavior analyses, although the additional analyses supported the direction of the findings. Third, differences across study cohorts may have influenced the estimates, given that UK Biobank participants are generally healthier than the general population,^49^ whereas FinnGen participants are comparatively less healthy due to differences in the recruitment procedures.^26^ Fourth, the focus on participants with European ancestry may limit the generalizability of these findings to other populations, while the differences in healthcare systems, expenditure structures, and patterns of healthcare utilization across countries must be considered when interpreting the broader implications and generalizability of the findings. Finally, the use of a dichotomous measure may limit the interpretation of the MR estimates for physical activity, particularly given the continuous nature of the underlying trait. Dichotomization may obscure variation across the exposure range, meaning that our results reflect the differences between the defined categories rather than changes in physical activity across its full spectrum.^31^

## Conclusions

The findings of this study extend the limited evidence regarding the causal effects of physical activity and sedentary behavior on healthcare costs. These results suggest that increasing physical activity and reducing sedentary behavior may reduce healthcare expenditure, potentially contributing to the long-term sustainability of both healthcare systems and society.

## Data Availability

All data produced in the present study are available upon reasonable request to the authors.

## Author Contributions

Mäkelä, Van Genechten, and Joensuu had full access to all of the data in the study and take responsibility for the integrity of the data and the accuracy of the data analysis.

*Concept and design*: All authors.

*Acquisition, analysis, or interpretation of data*: All authors.

*Drafting of the manuscript*: Mäkelä, Van Genechten, Kari, and Joensuu.

*Critical review of the manuscript for important intellectual content*: All authors.

*Statistical analysis*: Mäkelä and Van Genechten.

*Obtained funding:* Joensuu.

*Administrative, technical, or material support:* Joensuu.

*Supervision*: Bottas, Sillanpää, and Joensuu.

## Data availability statement

The data are publicly available from the GWAS catalog by study number, as described in Table 1.

## Ethics approval statement

This study relied on anonymous summary-level data that are publicly available. All the original studies had received ethical approval.

## Acknowledgements

We thank the personnel involved in collecting the original datasets for their valuable contributions. We are grateful to the authors of the original GWASs for their work and for making their data publicly available. We also thank the developers of the TwoSampleMR R package for providing their code and tools.

