## Supplement for "Causal Effects of Physical Activity and Sedentary Behavior on Healthcare Costs"

eFigure 1. Scatterplot for the causal relationship between self-reported MVPA and healthcare costs.
eFigure 2. Leave-one-out analysis for the self-reported MVPA
eFigure 3. Scatterplot for the causal relationship between device-based physical activity and healthcare costs.
eFigure 4. Leave-one-out analysis for the device-based PA
eFigure 5. Scatterplot for the causal relationship between self-reported sedentary behavior (Wang et al. 2022) and healthcare costs.
eFigure 6. Leave-one-out analysis for the self-reported sedentary behavior (Wang et al. 2022)
eFigure 7. Scatterplot for the causal relationship between self-reported sedentary behavior (van de Vegte et al. 2020) and healthcare costs.
eFigure 8. Leave-one-out analysis for the self-reported sedentary behavior (van de Vegte et al. 2020).
eFigure 9. Scatterplot for the causal relationship between device-based sedentary behavior and healthcare costs.
eFigure 10. Leave-one-out analysis for the device-based sedentary behavior instruments.

eTable 1. GWAS covariates

Genotyping, quality control and imputation

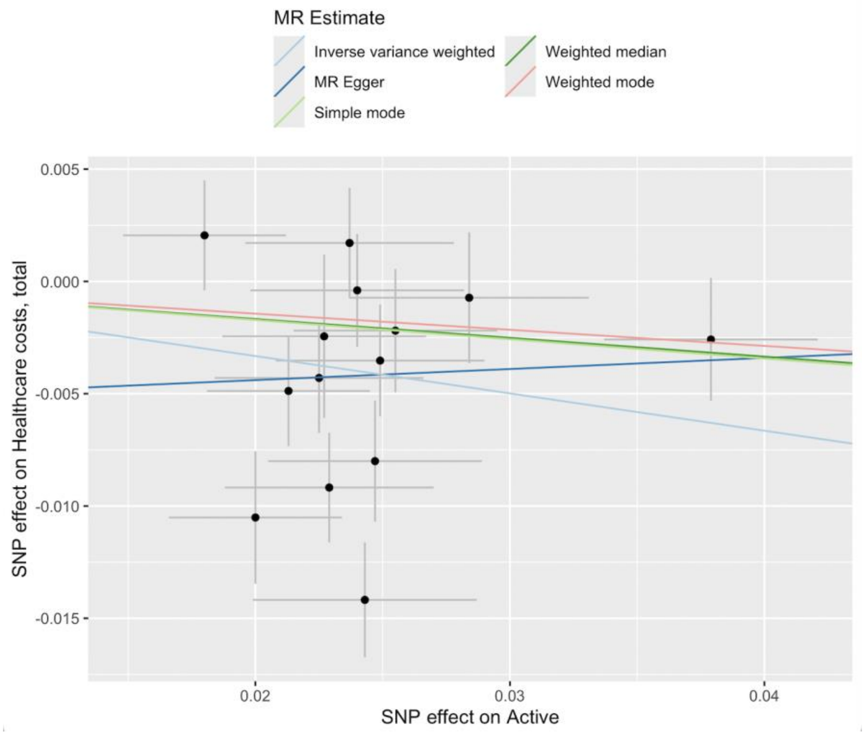


eFigure 1. Scatterplot for the causal relationship between self-reported MVPA and healthcare costs.

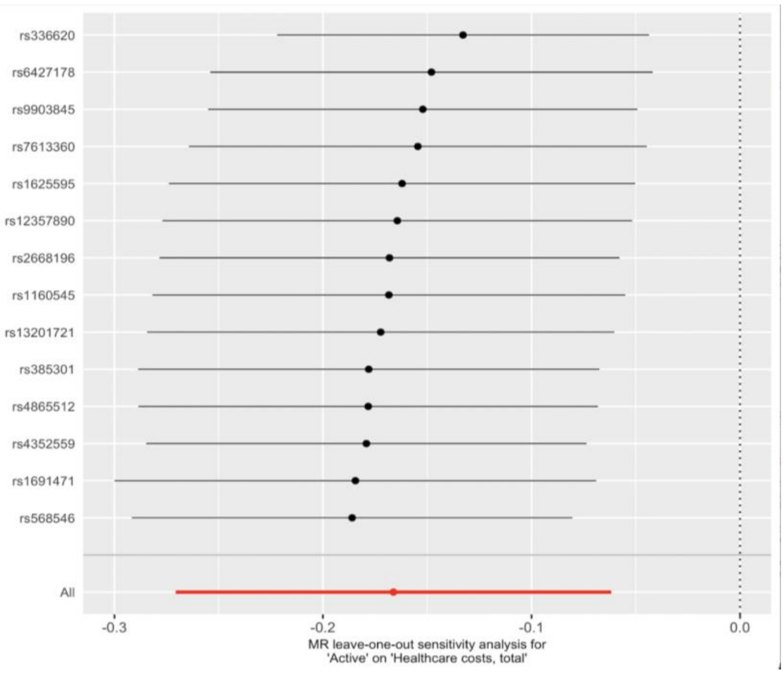


eFigure 2. Leave-one-out analysis for the self-reported MVPA

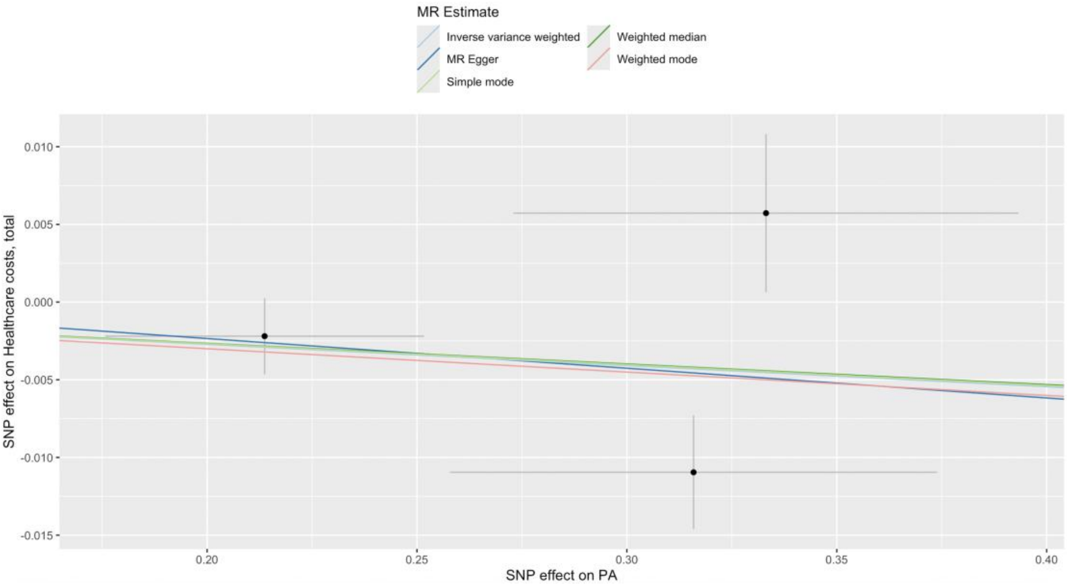


eFigure 3. Scatterplot for the causal relationship between device-based physical activity and healthcare costs.

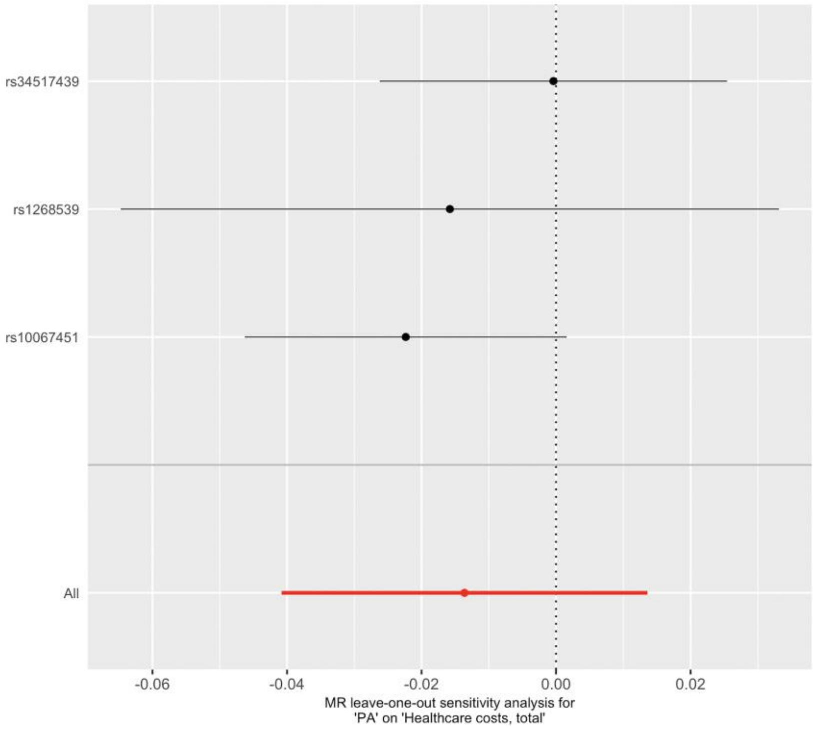


eFigure 4. Leave-one-out analysis for the device-based PA

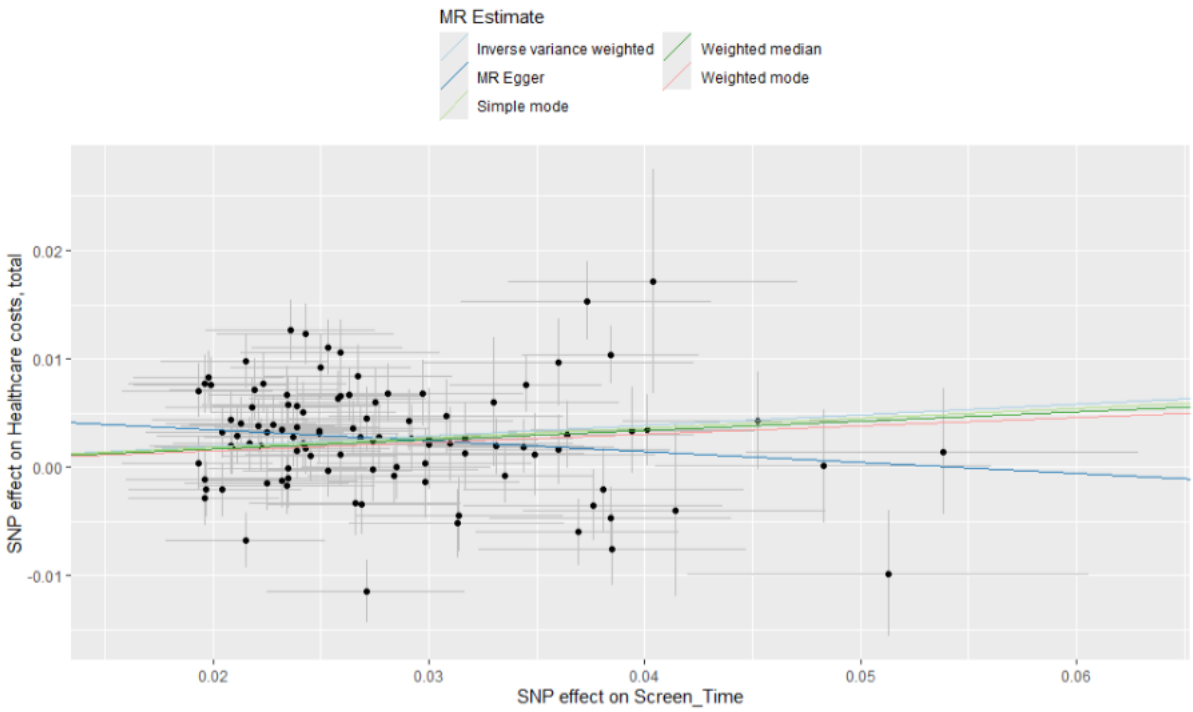


eFigure 5. Scatterplot for the causal relationship between self-reported sedentary behavior (Wang et al. 2022) and healthcare costs.

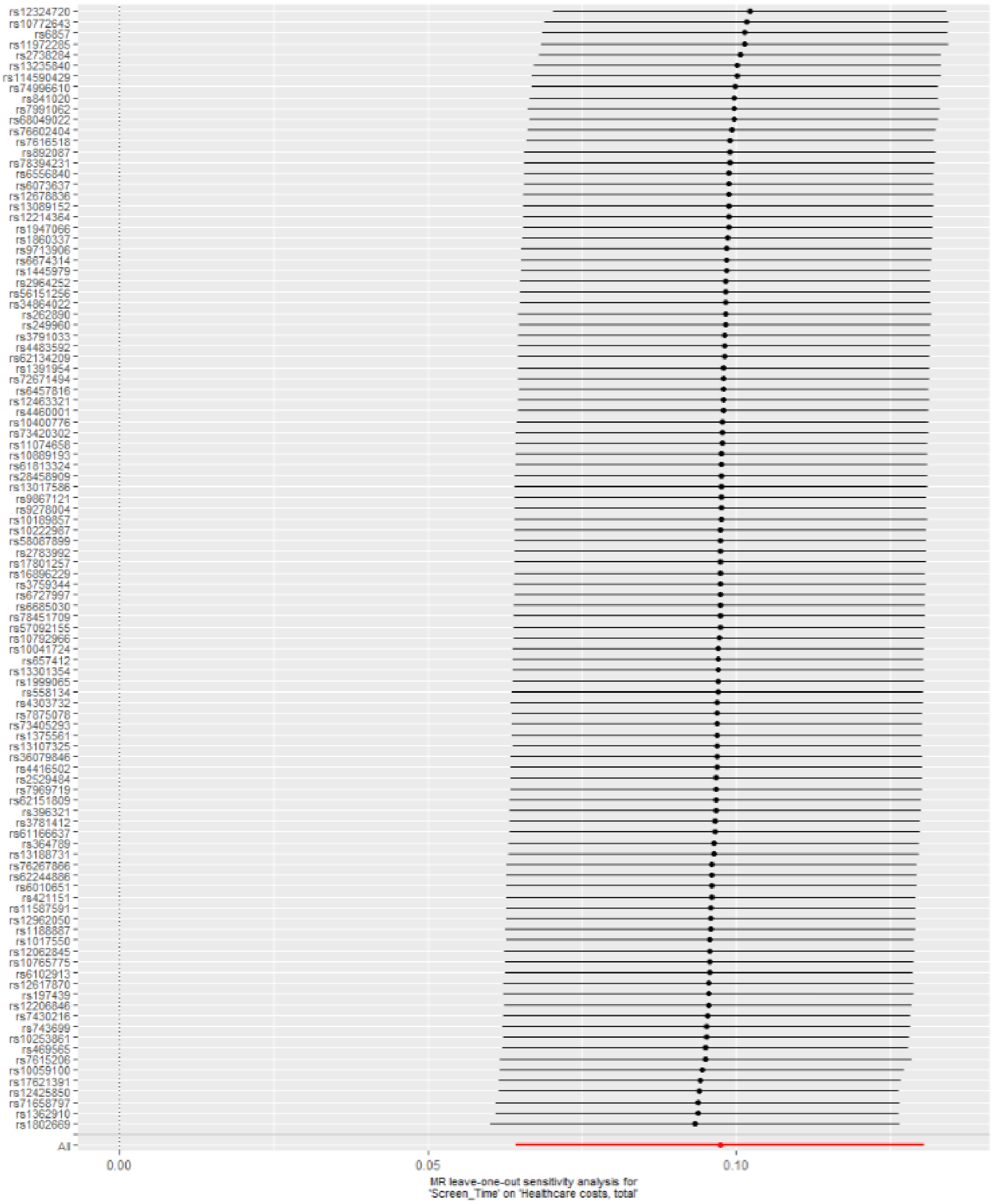

eFigure 6. Leave-one-out analysis for the self-reported sedentary behavior (Wang et al. 2022)

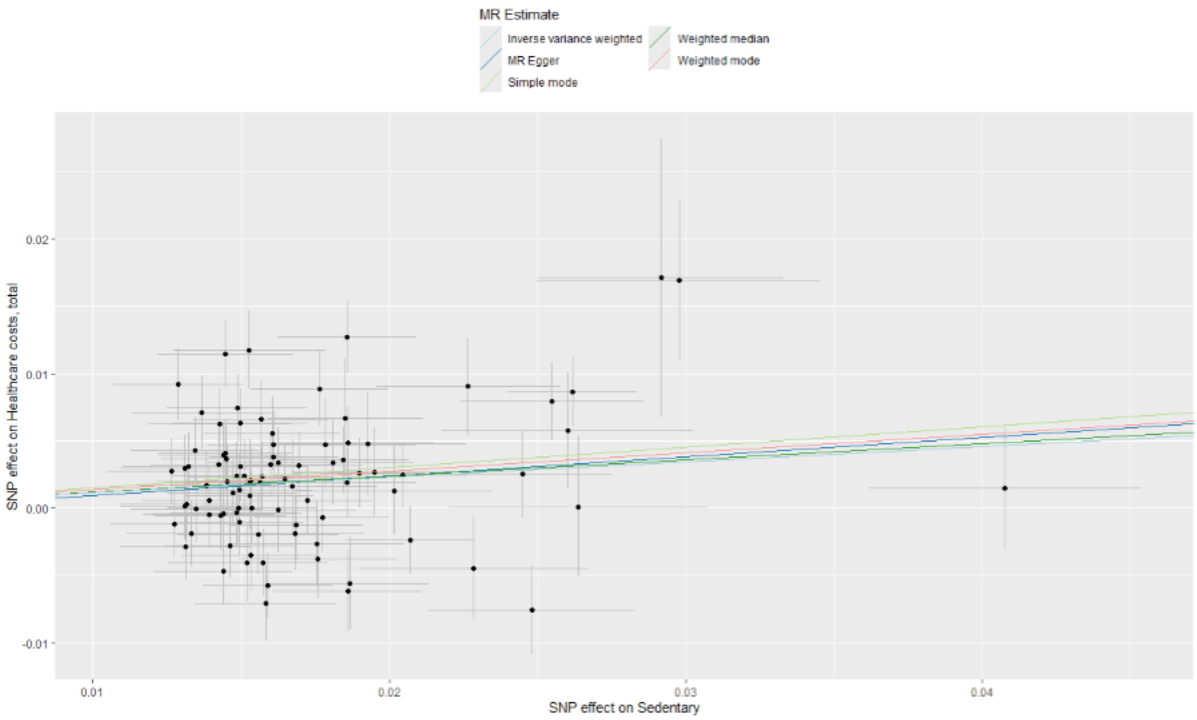


eFigure 7. Scatterplot for the causal relationship between self-reported sedentary behavior (van de Vegte et al. 2020) and healthcare costs.

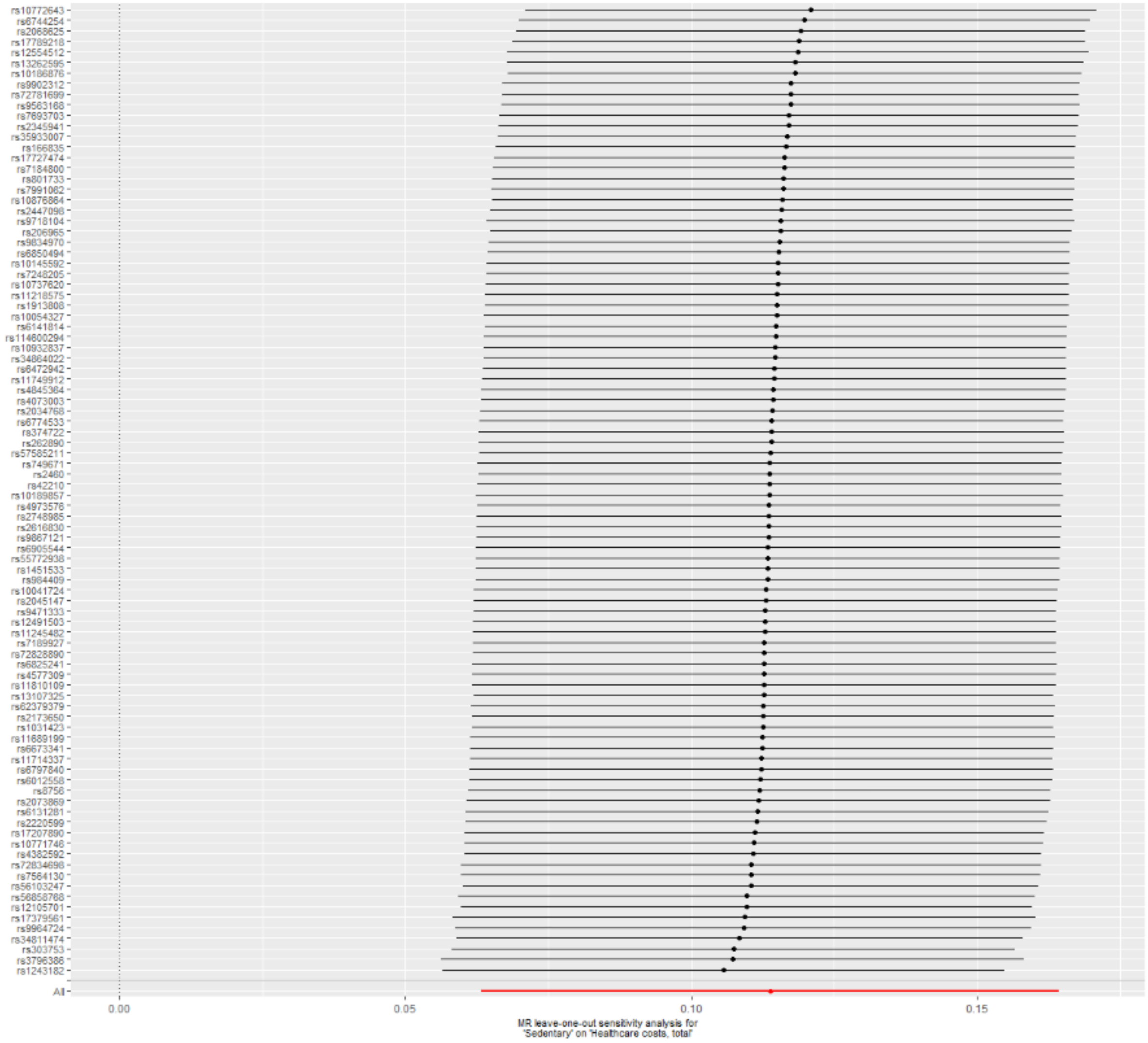


eFigure 8. Leave-one-out analysis for the self-reported sedentary behavior (van de Vegte et al. 2020).

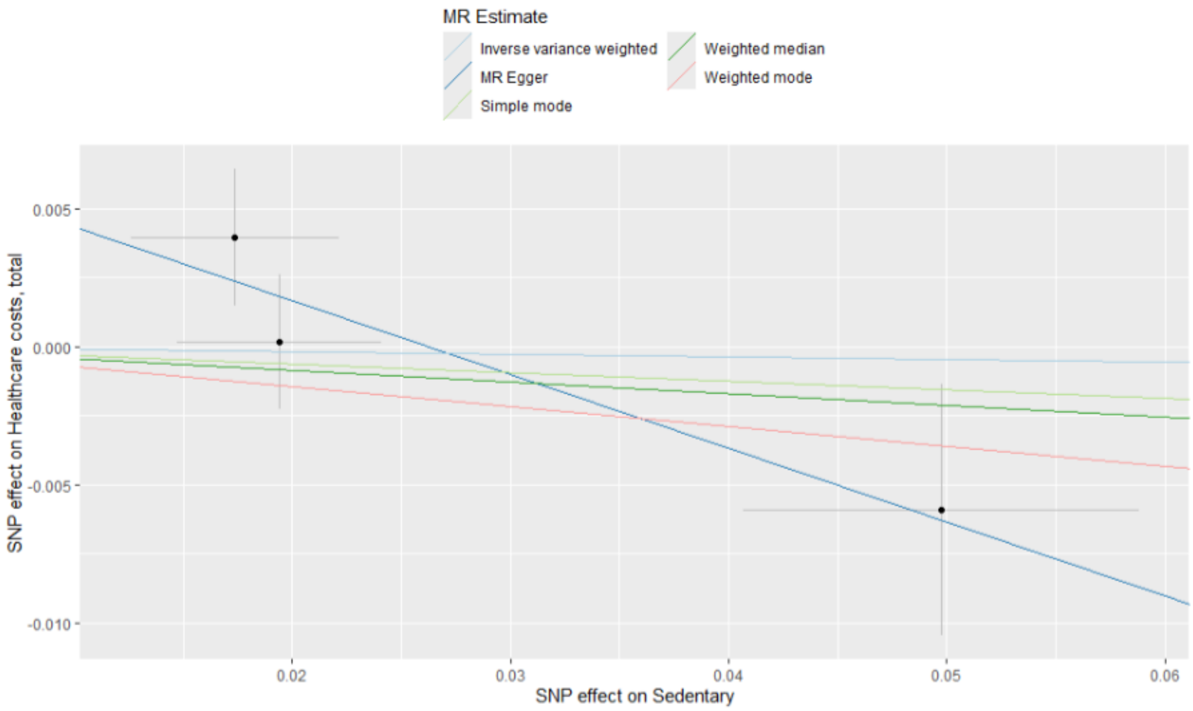


eFigure 9. Scatterplot for the causal relationship between device-based sedentary behavior and healthcare costs.

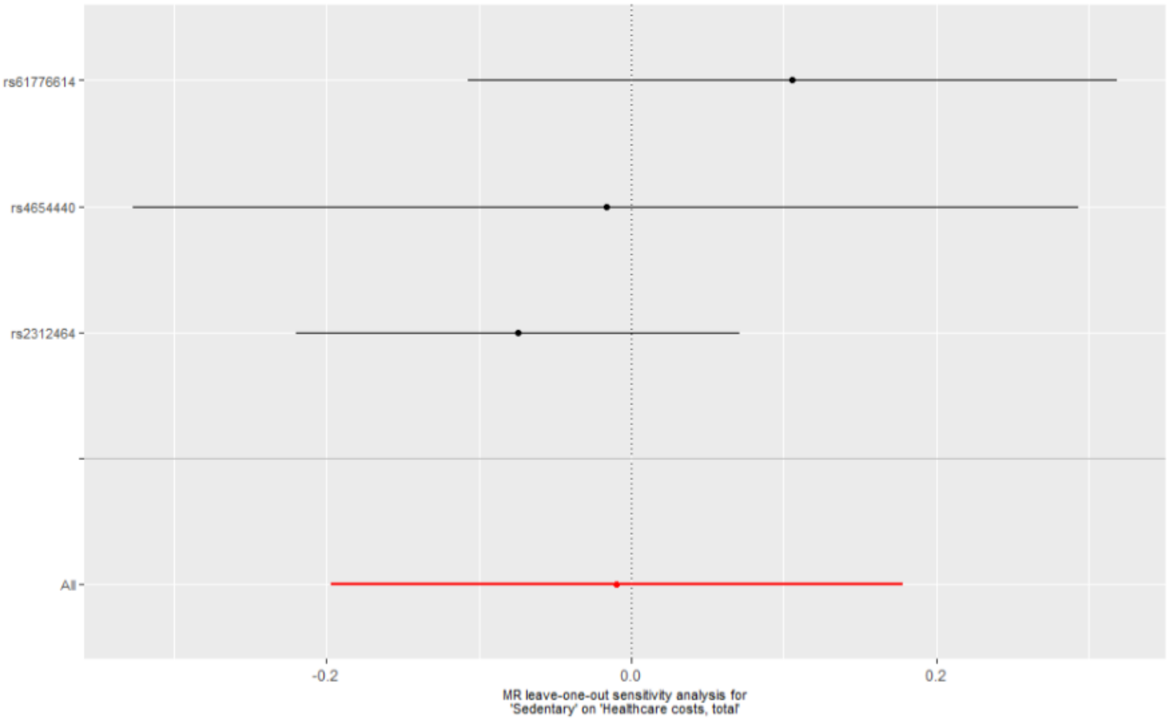


eFigure 10. Leave-one-out analysis for the device-based sedentary behavior instruments.

### eTable 1. GWAS covariates

| GWAS | Covariates |
| --- | --- |

| | |
|---|---|
| Self-reported MVPA and sedentary behaviour (Wang et al. 2022) | age, age-squared, principal components and additional study specific covariates.[1] |
| Self-reported sedentary behavior (van de Vegte et al. 2020) | age-squared, age, sex, age-sex interaction, the first 30 principal components. [2] |
| Device-based PA (Hanscombe et al. 2021) | age, age2, sex (for sex-combined analyses), array, centre and the first 10 principal components.[3] |
| Device-based SB (Doherty et al. 2018) | assessment centre, genotyping array, age, age squared, and season.[4] |
| Healthcare costs (Lee et al. 2023) | birth year, birth year squared, sex, 10 principal components, and batch covariates.[5] |

GWAS, Genome-wide association study: MVPA, moderate-to-vigorous physical activity; PA, physical activity, SB, sedentary behaviour.

**Genotyping, imputation and quality control**

Study specific protocols are described in detail in the original studies.[1–5]

Briefly, for Wang et al. (2022), genotyping platforms for the individual cohorts were ~60% Illumina-based, ~30% Affymetrix-based arrays, with the remainder using custom or mixed-platform designs. Genotype calling software was mainly GenomeStudio/BeadStudio/GenCall (Illumina) and Birdseed/BRLMM/Axiom GT1 (Affymetrix). Imputation softwares were predominantly MACH/Minimac and IMPUTE-family programs, with occasional use of BEAGLE, BIMBAM, and SHAPEIT. Quality control following study level analyses was conducted using standard procedures.[ref: 10.1038/nprot.2014.071]

For UK Biobank based studies (Doherty et al. 2018, van de Vegte et al. 2020, Hanscombe et al 2021), participants were genotyped using the UK Biobank Axiom Array and UK BiLEVE Axiom Array. Centralized quality control was applied at both the sample and variant levels, including checks for call rate, heterozygosity, sex discordance, relatedness, and population structure. Genotypes were phased and imputed using the Haplotype Reference Consortium (HRC) and the UK10K + 1000 Genomes reference panels [ref: Bycroft et al.].

The FinnGen individuals were genotyped with Illumina and Affymetrix chip arrays (Illumina Inc., San Diego, and Thermo Fisher Scientific, Santa Clara, CA, USA). For detailed information on genotyping, quality control, and imputation, please see the FinnGen website (https://finngen.gitbook.io/documentation/methods/genotype-imputation.
